# IMMUNE PROFILES TO DISTINGUISH HOSPITALIZED VERSUS AMBULATORY COVID-19 CASES IN OLDER PATIENTS

**DOI:** 10.1101/2022.06.23.22276820

**Authors:** Jéromine Klingler, Gregory S. Lambert, Juan C. Bandres, Rozita Emami-Gorizi, Arthur Nádas, Kasopefoluwa Y. Oguntuyo, Fatima Amanat, PARIS Study Team, Viviana Simon, Benhur Lee, Susan Zolla-Pazner, Chitra Upadhyay, Catarina E. Hioe

## Abstract

**Background:** A fraction of COVID-19 patients develops severe disease requiring hospitalization, while the majority, including high-risk individuals, experience mild symptoms. Severe disease has been associated with higher levels of antibodies and inflammatory cytokines, but the association has often resulted from comparison of patients with diverse demographics and comorbidity status. This study examined patients with defined demographic risk factors for severe COVID-19 who developed mild vs. severe COVID-19.

**Methods:** This study evaluated hospitalized vs. ambulatory COVID-19 patients in the James J. Peters VA Medical Center, Bronx, NY. This cohort presented demographic risk factors for severe COVID-19: median age of 63, >80% male, >85% black and/or Hispanic. Sera were collected four to 243 days after symptom onset and evaluated for binding and functional antibodies as well as 48 cytokines/chemokines.

**Findings:** Ambulatory and hospitalized patients showed no difference in SARS-CoV-2-specific antibody levels and functions. However, a strong correlation between anti-S2 antibody levels and the other antibody parameters was observed in hospitalized but not in ambulatory cases. Cytokine/chemokine levels also revealed differences, with notably higher IL-27 levels in hospitalized patients. Hence, among the older, mostly male patients studied here, SARS-CoV-2-specific antibody levels and functions did not distinguish hospitalized and ambulatory cases but a discordance in S2-specific antibody responses was noted in ambulatory patients, and elevated levels of specific cytokines were maintained in convalescent sera of hospitalized cases.

**Interpretation:** The data indicate that antibodies against the relatively conserved S2 spike subunit and immunoregulatory cytokines such as IL-27 are potential immune determinants of COVID-19.

**Research in context:** *Evidence before this study:* Previous studies demonstrated that high levels of SARS-CoV-2 spike binding antibodies and neutralizing antibodies were associated with COVID-19 disease severity. However, the comparisons were often made without considering demographics and comorbidities. Correlation was similarly shown between severe disease and marked elevation of several plasma cytokines but again, most analyses of cytokine responses to COVID-19 were conducted by comparison of patient cohorts with diverse demographic characteristics and risk factors.

*Added value of this study:* We evaluated here a comprehensive profile of SARS-CoV-2-specific antibodies (total Ig, isotypes/subtypes, Fab- and Fc-mediated functions) and a panel of 48 cytokines and chemokines in serum samples from a cohort of SARS-CoV-2-infected patients with demographic risk factors for severe COVID-19: 81% were male, 79% were >50 years old (median of 63), and 85% belonged to US minority groups (black and/or Hispanic). Comparison of hospitalized vs. ambulatory patients within this cohort revealed two features that differed between severe vs. mild COVID-19 cases: a discordant Ab response to the S2 subunit of the viral spike protein in the mild cases and an elevated response of specific cytokines and chemokines, notably IL-27, in the severe cases.

*Implications of all the available evidence:* Data from the study identified key immunologic markers for severe vs. mild COVID-19 that provide a path forward for investigations of their roles in minimizing or augmenting disease severity.

## Background

The emergence and rapid spread of the severe acute respiratory syndrome coronavirus 2 (SARS-CoV-2) in Wuhan, China, in December 2019 has led to a pandemic that continues to impact people worldwide. While research about this virus has progressed rapidly, leading to expeditious development of many types of diagnostic tests, treatments, and vaccines, important questions remain about the dynamic virus-host interactions that result in a wide range of disparate disease outcomes.

Coronavirus disease 2019 (COVID-19), the disease caused by SARS-CoV-2, can occur with different clinical manifestations. During the initial outbreak in China, most patients presented with mild to moderate symptoms which resolved without medical interventions, but ∼15% of patients progressed rapidly to severe disease requiring hospitalization (1,2). Among the hospitalized, patients also require different levels of intervention. Around a third of hospitalized patients developed acute respiratory disease syndrome (ARDS) and require mechanical ventilation (3). Elderly patients and individuals with comorbidities, such as cardiovascular disease, diabetes mellitus, chronic lung disease, chronic kidney disease, obesity, hypertension, and cancer, have a much higher mortality rate than healthy younger adults (4). In addition, the overall COVID-19 case-fatality ratio is at least 2.4 times higher in men than in women (5–7).

Understanding the immune responses associated with disease severity and recovery is essential to develop and apply effective treatments against COVID-19. While high levels of binding antibodies (Abs), in particular IgA, and neutralizing Abs have been associated with disease severity (8–10), comparison of severe and mild cases was often without consideration for age, sex, and comorbidities. Indeed, a study of infected patients from a wide range of age groups (1-102 years) demonstrated that anti-SARS-CoV-2 Ab levels varied depending on age (11). The levels in the 1-10 years old group were around three- to four-fold higher than the 25-102 years old groups. Another study comparing SARS-CoV-2-infected male and female patients from diverse ethnic groups showed no difference in Ab titers (12). However, male patients had higher plasma levels of innate immune cytokines and more robust induction of non-classical monocytes, while female patients developed significantly more robust T cell activation (12). IgG Fc glycome composition was also shown to predict disease severity but comparison was within a patient cohort that included both sexes and had an age range of 21 to 100 years old (13).

In addition to Abs, cytokine responses in severe vs. mild COVID-19 cases have been evaluated. An association was shown between SARS-CoV-2 infection and marked elevation of several plasma cytokines including interferon gamma-induced protein 10 (IP-10), chemokine (C-X-C motif) ligand 16 (CXCL16), interleukin (IL)-1β, IL-2R, IL-4, IL-6, IL-8, IL-10 and IL-17 (10,14–21). Several other studies provided evidence that type I interferon deficiency may lead to severe COVID-19, implying that disease severity may be due to impaired viral clearance and uncontrolled viral replication due to poor induction of early innate immunity (15,22–26). Another study revealed a positive association of the illness duration in severe cases with levels of IL-8 and soluble IL-2Rα (27). Further, a longitudinal study on 40 hospitalized COVID-19 patients found 22 cytokines that were correlated with disease severity (28). Yet, in a study looking at cytokine and leukocyte profile of COVID-19 patients >60 vs. <60 years old, another set of cytokines/chemokines was found to correlate with older age, longer hospitalization, and a more severe form of the disease (29). Similar to the Ab studies, most analyses of cytokine responses to COVID-19 were conducted by comparison of patient cohorts with diverse demographics and risk factors.

Further investigations of Ab and cytokine responses to SARS-CoV-2 among racial/ethnic minority US populations at risk of developing severe COVID-19 are warranted. Indeed, a study among US veterans showed that black and Hispanic individuals have experienced an excess burden of SARS-CoV-2 infection that was not entirely explained by underlying medical conditions or where they lived or received care (30). Immune responses to vaccinations also are influenced by host genetic and demographic variables such as race, ethnicity, and sex, as demonstrated by induction of higher neutralizing Ab titers following rubella vaccination in individuals of African descent as compared to the European and Hispanic subjects, although the cytokine responses were comparable (31).

Here, we evaluated SARS-CoV-2-specific (spike, RBD, S1, S2, nucleoprotein) Ab responses and conducted multiplex analysis of cytokines and chemokines in a cohort with risk factors for severe COVID-19. Of the 52 VA subjects from James J. Peters VA Medical Center (JJP VAMC), 81% were male, 79% were >50 years old (median 63), and 85% belonged to US minority groups (black and/or Hispanic). All were infected with SARS-CoV-2, as confirmed by diagnostic RT-PCR, during April-November 2020. Convalescent serum samples collected four to 243 days post disease onset were studied. We compared different categories of COVID-19 patients within this VA cohort: ambulatory (n=42) vs. hospitalized (n=10) patients, COVID-19 patients with (n=24) vs. without (n=28) comorbidities. We also compared a subset of specimens (n=20) from hospitalized vs. ambulatory cases that were matched based on sex, time post-infection, comorbidities, and spike-specific Ig levels. In addition, pre-pandemic and contemporaneous COVID-19-negative samples were studied. The data demonstrate that, in this cohort of older and mostly male VA patients, hospitalized and ambulatory patients had comparable binding and functional Abs, but diverged in their responses to the S2 spike subunit. Moreover, heightened levels of certain cytokines were detected and maintained in convalescent sera from hospitalized vs. ambulatory cases.

## Methods

### Recombinant proteins

SARS-CoV-2 spike (full-length external region, amino acids 1-1213) and RBD (amino acids 319-541) proteins were produced as described before (32,33). S1 (amino acids 16-685), S2 (amino acids 686-1213), and nucleoprotein (amino acids 1-419) antigens were purchased from ProSci Inc, CA (catalog #97-087, #97-079 and #97-085, respectively). All antigens were of SARS-CoV-2 Wuhan-Hu-1 (WA1) strain.

### Human samples

Fifty-two COVID-19-convalescent sera samples and 49 COVID-19-negative samples (38 contemporaneous and 11 pre-pandemic) were collected at JJP VAMC under IRB#BAN-1604 and the JJP VAMC Quality Improvement project “Evaluation of the clinical significance of two COVID-19 serologic assays”. In addition, five SARS-CoV-2 seronegative serum samples were sourced from the PARIS cohort, which follows health care workers since March 2020 (IRB-20-03374, approved by the Mount Sinai Hospital Institutional Review Board). All participants in the PARIS cohort and other research protocols provided written informed consent and agreed to future research and sample sharing. Samples were coded prior to processing, testing, and sharing. Of the COVID-19-negative specimens, 21 were used in the Ab binding and activities experiments, and 28 were used for the cytokines induction experiments. Before use, all sera were heat-inactivated (30 min at 56 °C, for Ab binding and activities experiments) or treated with 1% Triton X-100 (30 min at room temperature, for the cytokines induction experiments).

### Ab binding assay

The initial Ab evaluation was done on the Abbott Architect instrument using the Abbott SARS-CoV-2 IgG assay, a chemiluminescent microparticle immunoassay to detect IgG against the virus nucleoprotein. Subsequent Ab analyses were performed using the multiplex bead Luminex platform, in which recombinant SARS-CoV-2 spike, RBD, S1, S2, and nucleoprotein antigens were coupled to beads and experiments performed as described in (34,35).

### Neutralization

SARS-CoV-2 pseudotyped particles (COV2pp) with spike protein of WA1 strain were produced and used in neutralization assays as described before (36–38).

### ADCP

Assays to measure spike-specific ADCP were performed using a published protocol (39) with some modifications reported elsewhere (35).

### Cytokines induction

The samples were tested for 48 cytokines/chemokines by Eve Technologies Corporation, Canada (Human Cytokine/Chemokine 48-Plex Discovery Assay® Array (HD48)). Data were presented as concentrations (pg/mL) or modified Z scores. The modified Z scores used to normalize the data were calculated as:

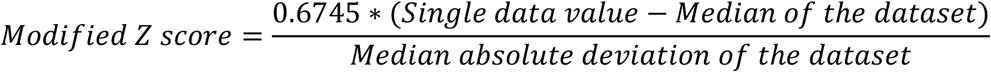

### Statistical analysis

Statistical analyses were performed as designated in the figure legends using GraphPad Prism 9 (GraphPad Software, San Diego, CA).

## Results

### Ab responses to different SARS-CoV-2 antigens among hospitalized and ambulatory VA patients

Fifty-two COVID-19-convalescent (**Table 1**) and 21 COVID-19-negative serum samples were collected from a cohort of patients who received care at the JJP VAMC. Samples were collected between March and December 2020, during the first year of COVID-19 pandemic in New York City. COVID-19 diagnosis was confirmed by positive PCR test. The vast majority of the COVID-19-convalescent subjects was male (80%), Black/Hispanic (85%), and >50 years old (79%, median=63, Q1-Q3=52-74), which are representative demographics of the US Veterans population. The preponderance for male and older age in this cohort allowed for characterization of immune responses in the population known to be at risk for severe COVID-19. The COVID-19 patients were categorized in two groups: hospitalized (n=10) vs. ambulatory (n=42). No difference in age and race/ethnicity was observed between the hospitalized vs. ambulatory patients (**Fig 1a**). However, all the hospitalized patients were male, while 10 of the 42 ambulatory patients were female. Analyses also considered the presence or absence of comorbidities known to be associated with more severe COVID-19, which included HIV, obesity, chronic obstructive pulmonary disease, diabetes, asthma, end stage renal disease or pulmonary embolism.

**Fig 1.**
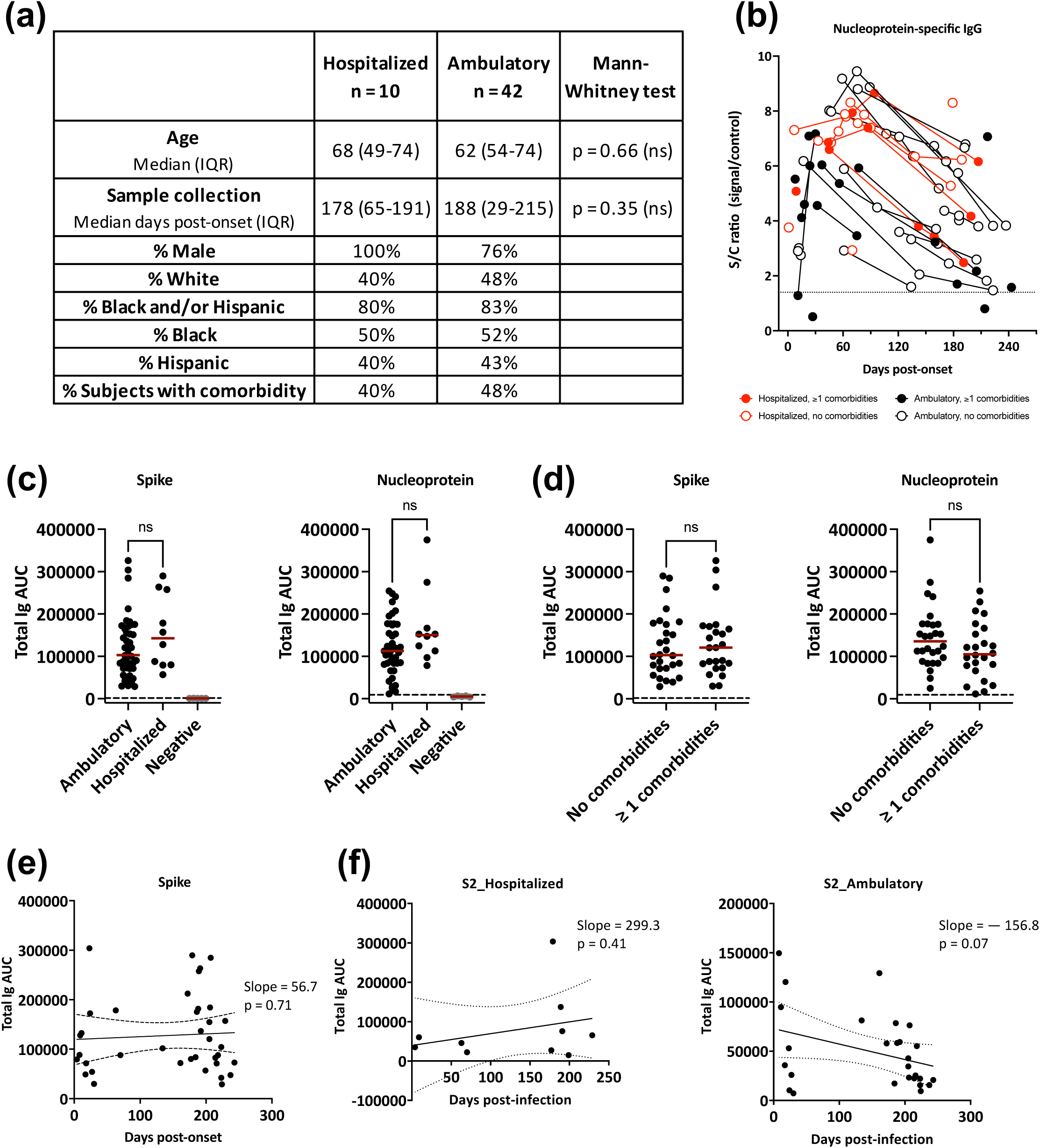
Total Ig antibody responses to SARS-CoV-2 among hospitalized and ambulatory patients in the VA cohort. **a**. Demographic and clinical data of hospitalized (n = 10) vs. ambulatory (n = 42) patients. **b**. IgG responses to nucleoprotein in hospitalized and ambulatory patients with or without comorbidities. The dotted line represents the cut-off at 1.4. **c**. Total Ig levels against spike and nucleoprotein in hospitalized (n = 10) vs. ambulatory (n = 42) patients. AUC: area under the curve. The red line represents the median. The dotted line represents the cut-off, calculated as mean + 3SD of 5 COVID-19-negative specimens. **d**. Total Ig levels against spike and nucleoprotein between patients with (n = 24) vs without (n = 28) comorbidities. The red line represents the median. The dotted line represents the cut-off, calculated as mean + 3SD of 5 COVID-19-negative specimens. **e**. Changes of spike-specific total Ig levels over time post-symptom onset. The dotted lines represent the linear regression 95% confidence bands. **f**. Changes of S2-specific total Ig levels over time post-symptom onset in hospitalized (n = 10, left) vs. ambulatory (n = 42, right) patients. The dotted lines represent the linear regression 95% confidence bands.

Longitudinal specimens of the COVID-19 patients were initially tested for SARS-CoV-2 nucleoprotein-specific IgG (**Fig 1b**). These COVID-19 patients generated IgG responses against nucleoprotein, which peaked around day 75 post-infection. The responses declined subsequently, but the Abs were detected above cut-off up to 250 days post-infection in all except for two ambulatory patients. Samples from these two patients, collected at day 27 and day 214 post-infection, although negative for nucleoprotein-specific IgG (**Fig 1b**), were positive for spike-specific IgM and IgG, respectively (data not shown) and were both positive for total Ig against spike and nucleoprotein (**Fig 1c**). Of note, the anti-nucleoprotein IgG levels among hospitalized patients overlapped with those of ambulatory patients, and no difference was apparent in the peak levels and the decline rates between the two groups (**Fig 1b**). There was also no apparent clustering of patients with vs. without comorbidities (**Fig. 1b**).

The specimens collected at the last time point from each subject were available for further investigation. The time points ranged from 4-243 days after symptom onset, with median of 178 and 188 days for hospitalized and ambulatory group, respectively (**Fig 1a**). To examine the relative levels of Abs induced by hospitalized vs. ambulatory patients against spike and its domains as compared to nucleoprotein, the serum samples were titrated for total Ig against spike, RBD, S1, S2 and nucleoprotein antigens (**Fig 1c-e and Supplemental Fig 1**). COVID-19 patients induced highly variable levels of Abs against each of the five antigens tested, but all displayed Ig reactivity above cut-off (calculated as mean + three standard deviations (SD) of five COVID-19-negative specimens) (**Fig 1c and Supplemental Fig 1a**). The S2-specific Ab levels were relatively low (**Supplemental Fig 1a**), similar to past reports on other cohorts of infected and vaccinated subjects (36). There was a trend of higher median levels of Abs against all five antigens in the hospitalized group as compared to the ambulatory group, but the differences did not reach statistical significance and the individual Ab levels from the two groups essentially overlapped (**Fig 1c and Supplemental Fig 1a**), indicating that the two groups of patients could not be differentiated by their anti-SARS-CoV-2 Ab levels. Moreover, there was no difference in the Ab levels of patients with vs. without comorbidities (**Fig 1d and Supplemental Fig 1b**).

Because the sample collection times were over a wide range of days after disease onset, we searched for changes in total Ig levels against each of the five SARS-CoV-2 antigens over time after disease onset (**Fig 1e and Supplemental Fig 1c**). The Ig responses to spike did not decline over the 250 days post-onset (slope=56.7 and p=0.71) (**Fig 1e**). Ig levels against RBD and S1 also did not decrease over this period, while the S2- and nucleoprotein-specific Ig levels demonstrated a non-significant decline (**Supplemental Fig 1c**). Interestingly, when the specimens were divided according to the disease severity, the S2-specific Ig levels overtime showed a trend of positive slope for the hospitalized patients and negative slope for the ambulatory patients (**Fig 1f**). This pattern was not seen with Abs against S1 or RBD (data not shown).

### Neutralization activities against SARS-CoV-2 among hospitalized and ambulatory VA patients

COVID-19-convalescent sera were tested for neutralization activities against SARS-CoV-2 using a pseudovirus bearing SARS-CoV-2 spike protein (WA1 strain) as performed previously (36–38). COVID-19-negative sera (n=21) were tested in parallel as control. Samples from all COVID-19-positive patients had neutralization activity reaching readily above 50%, while none of the COVID-19-negative sera did (IC_50_ <10) (**Fig 2**). Neutralization titers of hospitalized and ambulatory patients did not differ, although the sample size of the hospitalized group was small (n=10). The IC_50_ values ranged from 60 to 3313 for hospitalized patients (median=194) and from 43 to 4402 for ambulatory patients (median=162.5). Neutralization titers were also similar between groups with vs. without comorbidities associated with severe COVID-19 (**Fig 2b**). In addition, the neutralization titers showed no association with age (**Fig 2c**) and no decline with time (**Fig 2d**), demonstrating that these parameters do not influence SARS-CoV-2-neutralizing titers detected in sera of this VA cohort.

**Fig 2.**
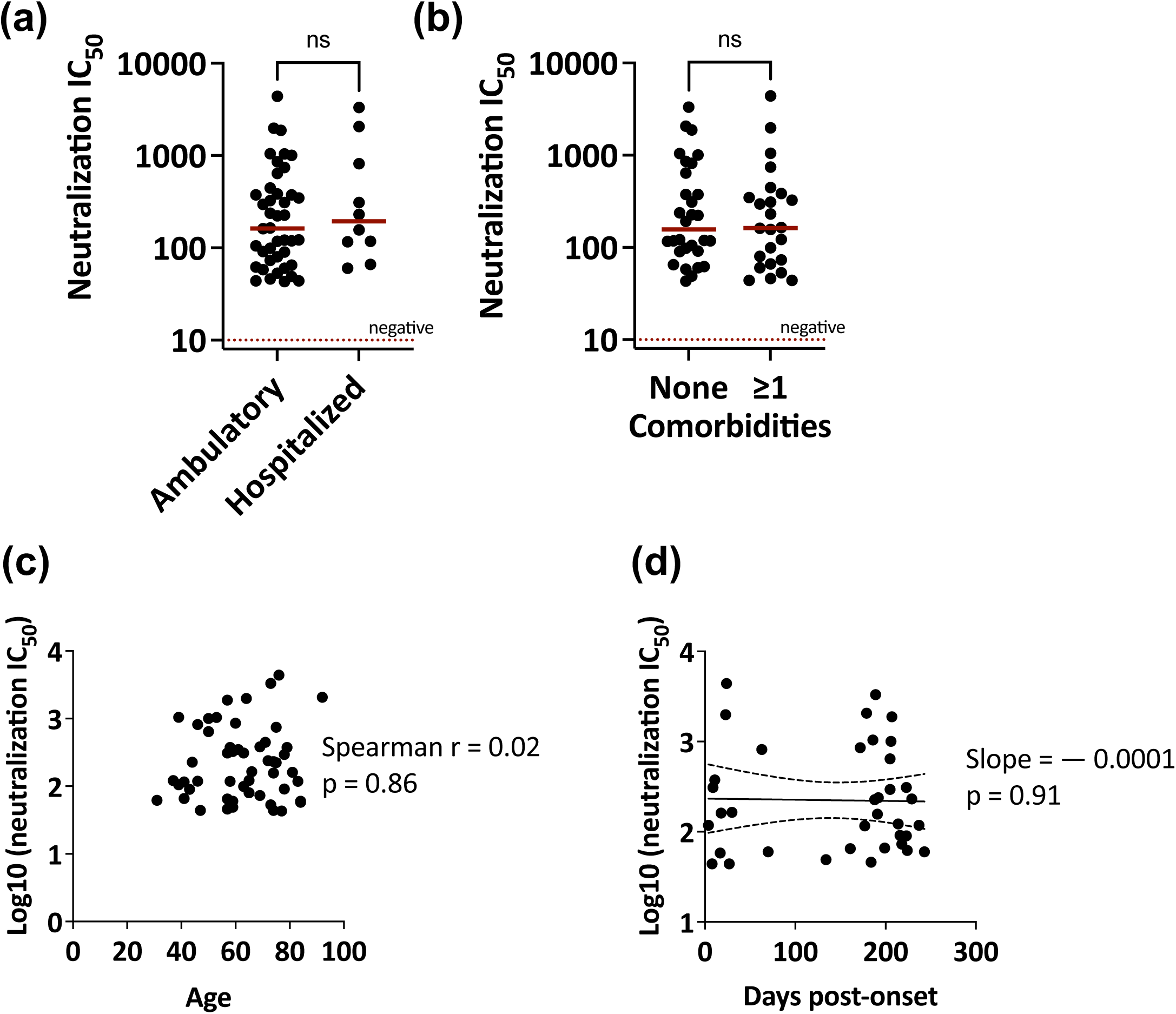
Neutralization activities against SARS-CoV-2 among hospitalized and ambulatory patients in the VA cohort. **a**. Neutralization titers in sera of ambulatory (n = 42) vs. hospitalized (n = 10) patients. The red line represents the median. Negative is set at 10, the lowest reciprocal dilution. **b**. Neutralization titers in sera of patients with (n = 24) vs. without (n = 31) comorbidities. The red line represents the median. Negative is set at 10, the lowest reciprocal dilution. **c**. Spearman correlation between neutralization titers and age of patients. **d**. Changes in neutralization titers over time post-symptom onset. The dotted lines represent the linear regression 95% confidence bands.

### Isotypes of serum Abs against SARS-CoV-2 spike produced by hospitalized and ambulatory patients

To understand the Fc properties of Abs raised against SARS-CoV-2 in patients presenting with distinct COVID-19 severity, we examined spike-specific Ig isotypes in a subset of hospitalized (n=6) vs. ambulatory cases (n=14) in which each hospitalized patient was matched to two or three ambulatory patients based on sex, time post-infection (early: <25 days vs. late: >130 days post-infection), comorbidities (none vs. at least one), and spike-specific total Ig levels (< half a log10) (**Table 2**). Four COVID-19-negative plasma specimens were included to establish background levels. All the patients had IgM, IgG1, IgG3 and IgA1 spike-specific Abs. The levels of these isotypes and subtypes did not differ between the hospitalized and ambulatory groups (**Fig 3**). Similar results were observed with Ig isotypes against RBD (**Supplemental Fig 2**). Some patients also mounted IgG2 and IgG4 Ab responses. The levels of these IgG subtypes were relatively low, and no difference was apparent between matched hospitalized and ambulatory patients. These data demonstrate that hospitalized and ambulatory patients display a comparable array of serum Ig isotypes against SARS-CoV-2 spike and RBD.

**Fig 3.**
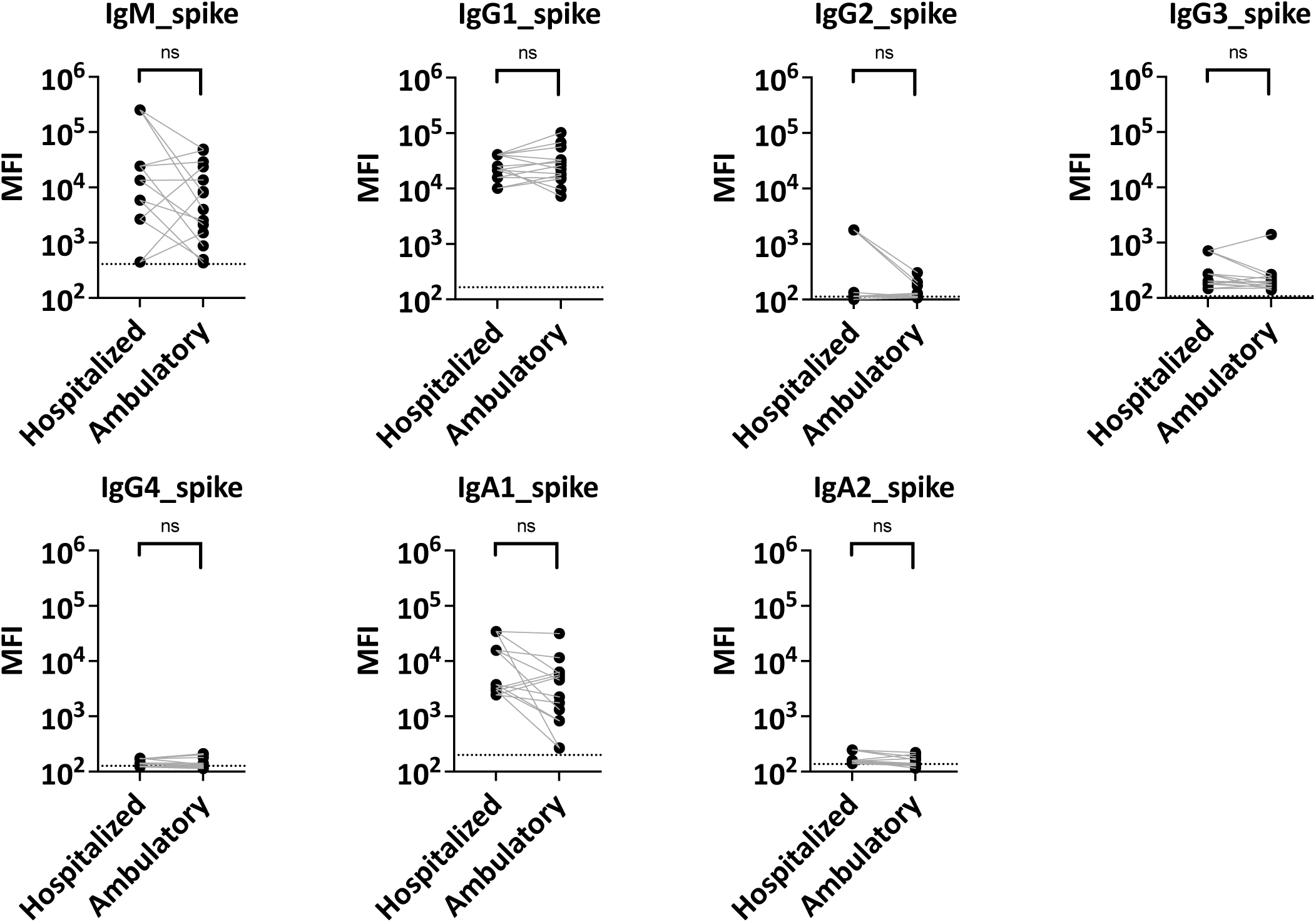
Antibody isotypes against SARS-CoV-2 in sera of matched hospitalized vs. ambulatory VA patients. The levels of spike-specific IgM, IgG1-4, IgA1 and IgA2 were measured in sera from matched hospitalized vs. ambulatory patients. The data are represented as mean fluorescent intensity (MFI) at a 1:200 serum dilution, each sample was tested in duplicate. The dotted line represents the cut-off. Statistical significance was calculated by a Wilcoxon matched-pairs test; each hospitalized patient was compared to 2 or 3 matched ambulatory patients.

### Fab-mediated neutralization and Fc-mediated activities of SARS-CoV-2-specific Abs among matched hospitalized and ambulatory VA patients

The matched set of hospitalized and ambulatory specimens was next tested for Ab activities against SARS-CoV-2. In addition to Fab-mediated neutralization, we examined Fc-mediates activities: spike-specific ADCP and spike- or RBD-complement binding (C1q and C3d) (**Fig 4 and Supplemental Fig 3**). Similar to the neutralization data shown in **Fig 2** for all samples, neutralization levels between matched hospitalized (median IC_50_ of 175) and ambulatory (median IC_50_ of 142) specimens were not different (**Fig 4a**). While certain hospitalized patients displayed higher IC_50_ than their matched ambulatory counterparts, some others had lower IC_50_ values. Fc-mediated spike-specific ADCP also showed no apparent trend, and the levels were indistinguishable between hospitalized and ambulatory groups (median AUC of 518 and 485, respectively) (**Fig 4b**). When complement binding and activation were examined, the capacity of spike- and RBD-specific Abs for C1q binding was found to be similar for both groups, and the C3d deposition levels also did not differ (**Fig 4c-d and Supplemental Fig 3**).

**Fig 4.**
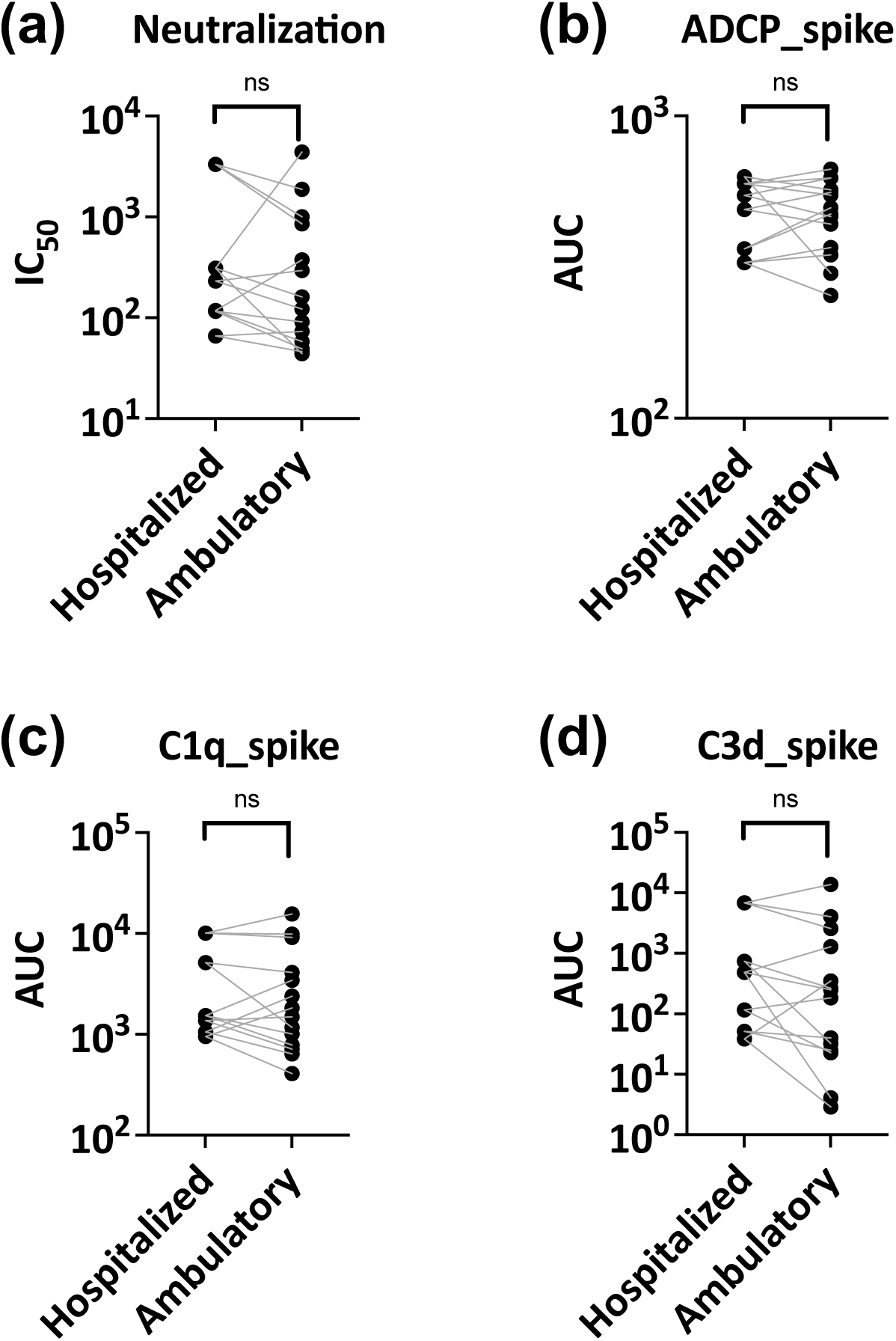
Fab-mediated neutralization and Fc-mediated activities of SARS-CoV-2-specific antibodies in sera from matched hospitalized and ambulatory VA patients. **a**. Neutralization activities in matched hospitalized vs. ambulatory patients. IC_50_.: reciprocal serum titers yielding 50% neutralization. **b**. Spike-specific ADCP activities in matched hospitalized vs. ambulatory patients. **c**. C1q binding to spike-specific antibodies in matched hospitalized vs. ambulatory patients. **d**. C3d binding to spike-specific antibodies in matched hospitalized vs. ambulatory patients. AUC: area under the titration curve. Statistical significance was calculated by a Wilcoxon matched-pairs test: six specimens from hospitalized patients were compared to the average of 2 or 3 matched specimens from ambulatory patients. ns: not significant (p >0.05).

Correlation matrix was compiled from both binding and functional Ab data of the hospitalized and matched ambulatory groups. A remarkable difference was noted: in the hospitalized group, a strong positive correlation was observed between the levels of S2-specific Ig and the other Ab parameters tested, whereas the correlation was weaker or absent in the ambulatory group (**Fig 5a-b**). Indeed, in the ambulatory group, poor correlation was apparent between the anti-S2 Ig levels and the Ig levels against the entire spike protein, S1, or RBD (**Fig 5b**). These results indicate a discordance of Ab responses against the relatively conserved S2 subunit vs. the rest of spike regions in the ambulatory cases but not the hospitalized cases.

**Fig 5.**
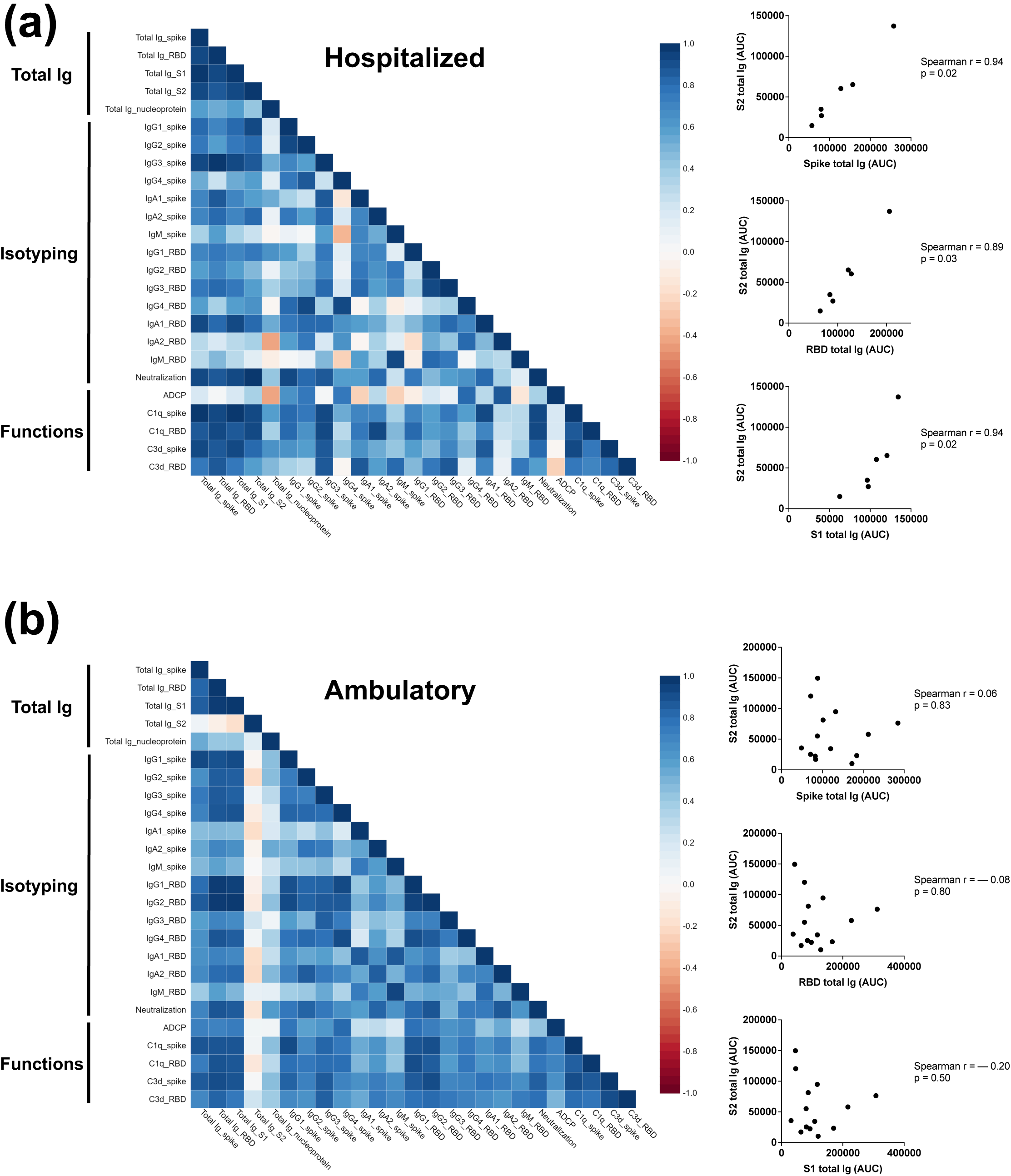
Correlations of binding and functional Ab levels in matched hospitalized and ambulatory VA patients. Spearman correlation matrix of serum Ab binding levels and functional activities were generated to compare matched **(a)** hospitalized (n = 6) vs. **(b)** ambulatory (n = 14) patients.

### Differences in cytokines responses among hospitalized and ambulatory patients

In addition to evaluating the Ab responses, we examined the cytokines profile of these convalescent COVID-19 patients. Fifty-two COVID-19-convalescent patients and 28 COVID-19 negative sera were tested for 48 cytokines and chemokines, many of which participate in induction and modulation of inflammatory responses (**Figs 6 and 7, Supplemental Figs 4-7**). Twelve cytokines/chemokines were found to be significantly different between COVID-19-positive and negative groups (**Fig 6a, Supplemental Fig 4**). MDC and RANTES were significantly higher in COVID-19 convalescent sera than negative controls, while the remaining 10 were lower in COVID-19 convalescent sera, with sCD40L and IL-17E/IL-25 displaying p values of <0.0001. Linear regression analysis of these 12 cytokines showed that the cytokine levels did not change significantly up to 250 days after COVID-19 disease onset (**Supplemental Fig 5**), except for IL-15 and M-CSF which declined over time (slope= −0.001 and −0.002, and p=0.02 and 0.007).

**Fig 6.**
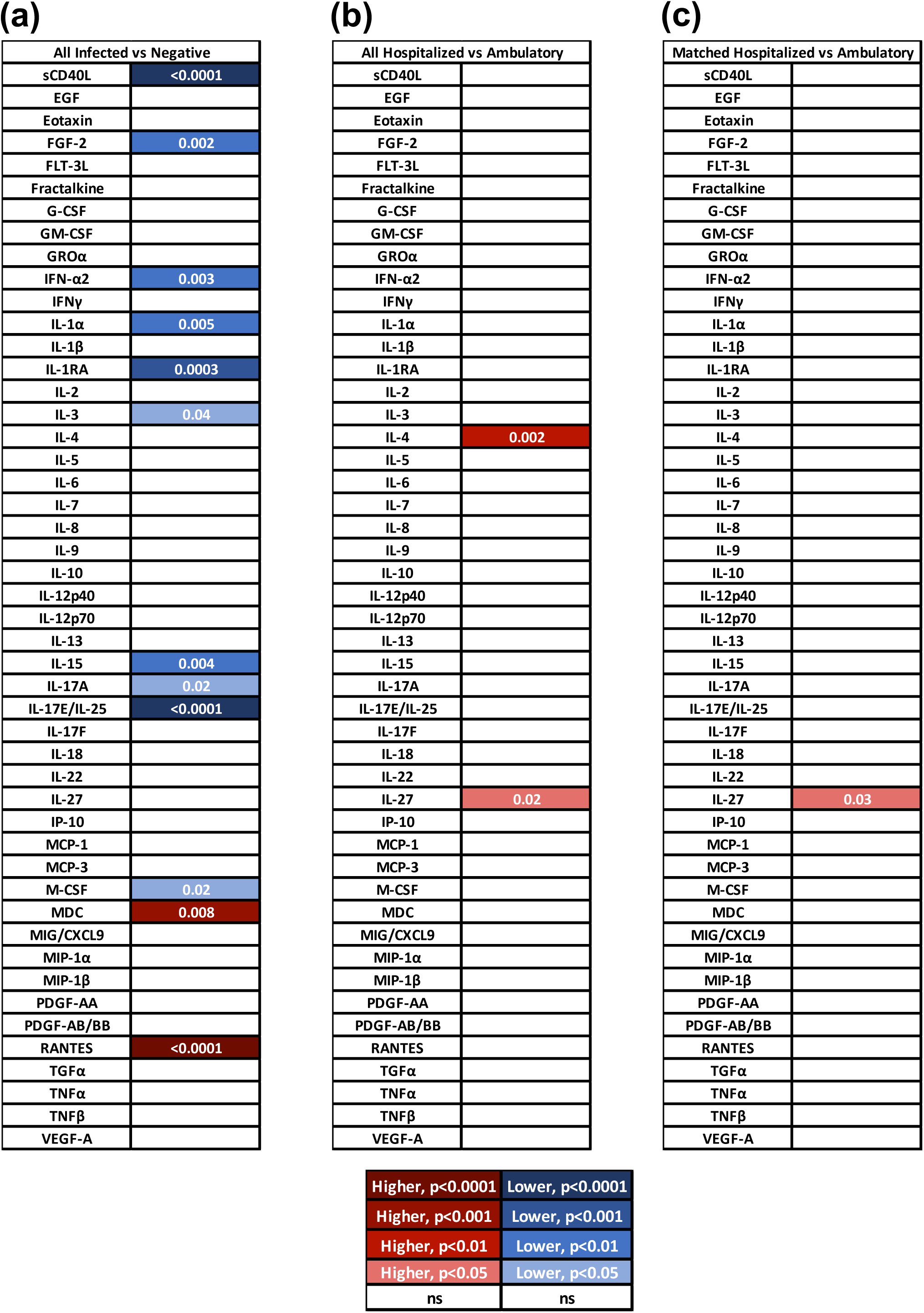
Differential cytokine/chemokine responses among hospitalized vs. ambulatory VA patients. Statistically different responses in infected vs. negative individuals **(a)**, hospitalized vs. ambulatory patients **(b)** and matched hospitalized vs. ambulatory patients **(c)** were indicated by color codes. Statistical significance was calculated by unpaired **(a, b)** or paired **(c)** non-parametric t-test. Significance was similarly achieved with other tests (parametric t-test and non-parametric Mann Whitney Wilcoxon in R).

**Fig 7.**
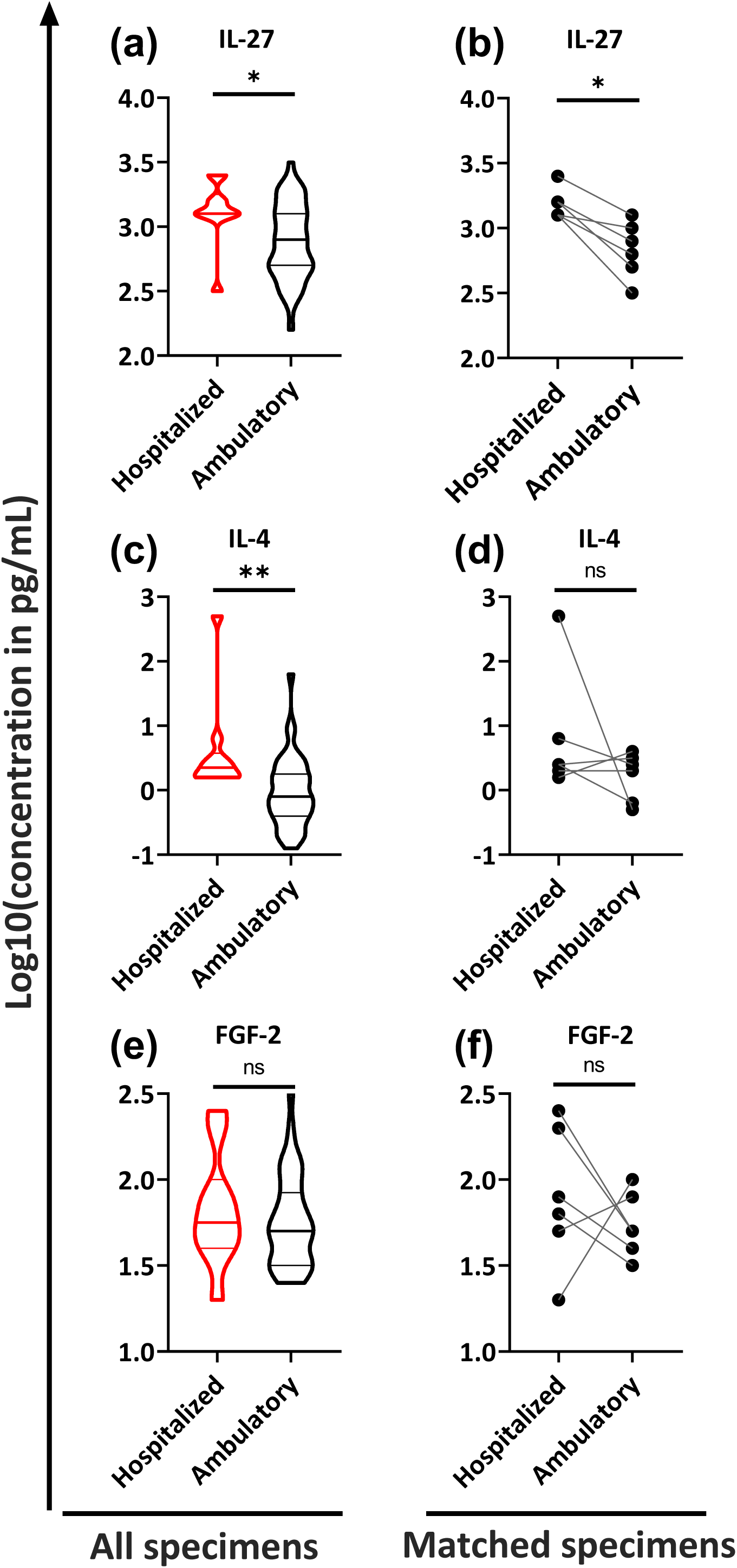
Cytokines responses among hospitalized and ambulatory VA patients. Data from three cytokines (IL-27, IL-4, and FGF-2 are presented as examples to show similarities and differences between hospitalized vs. all ambulatory patients **(a, c, e)** or between matched hospitalized vs. ambulatory patients **(b, d, f)**. Statistical significance was calculated in GraphPad Prism by unpaired **(a, c, e)** or paired **(b, d, f)** non-parametric t-test (*: p<0.05, **: p<0.01, ns: p>0.05). Significance was similarly achieved with parametric t-test and non-parametric Mann Whitney Wilcoxon in R.

We subsequently compared COVID-19 patients with different disease severity and asked whether the cytokines/chemokine responses were elevated in hospitalized patients compared to ambulatory patients (**Supplemental Figs 6b and 6c, and Fig 7**). The median modified Z scores showed an overall higher response in the hospitalized vs. ambulatory patients (p=0.03), with higher median values observed in a constellation of cytokines/chemokines (e.g. IL-4, IL-7, IL-8, IL-13, IL-17A, IL-17E/IL-25, IL-18, IL-22, IL-27, MCP-3, M-CSF, MDC, MIG/CXCL9, MIP-1α, MIP-1ß, PDGF-AA, and TNF-ß) (**Supplemental Fig 6b**). Correlation matrices of these cytokine/chemokine data further supported the notion of a stronger coordination of cytokine/chemokine alterations among the hospitalized patients as compared to the ambulatory patients whose correlation matrix was similar to that of the negative control (**Supplemental Fig 7**).

Comparison was also performed on individual cytokines or chemokines, and two cytokines were found to be significantly higher in hospitalized vs. ambulatory patients: IL-4 and IL-27 (**Fig 6b**). These were distinct from the 12 found to differ between COVID-19 positive and negative groups, and their levels did not significantly decline over 250 days post disease onset (slope= – 0.0001 and – 0.0005, and p >0.05, **Supplemental Fig 5**). When we evaluated the subset of matched severe and mild specimens, only IL-27 remained significantly higher in hospitalized vs. ambulatory patients with p value of 0.03 (**Fig 6c**). Therefore, the higher IL-27 levels between the hospitalized vs. ambulatory groups were evident in the plots of all COVID-19 patients (**Fig 7a**) and in the matched subset (**Fig 7b**). For comparison, two other cytokines were also plotted (**Fig 7c-f**). IL-4 was higher in sera of hospitalized vs. all ambulatory patients (**Fig 7c**) but showed inconsistent patterns in the matched subsets (**Fig 7d**). FGF-2, on the other hand, was comparable in hospitalized patients and all or matched ambulatory patients (**Fig 7e, f**). Notably, the IL-27 levels correlated with IL-1α, IL-3, IL-6 and TNF-α (Spearman r=0.61–0.75 and p=0.01–0.03) in the hospitalized patients, but not in the ambulatory patients (**Supplemental Fig 7**).

## Discussion

This study examined serum Ab and cytokine/chemokine responses in SARS-CoV-2-infected patients with demographic risk factors for developing severe COVID-19, which included older age, male sex, and Black/Hispanic racial or ethnic background. Most patients (81%), however, had mild disease requiring no hospitalization, and only ten were hospitalized. Comparison of the hospitalized vs. ambulatory patients within the cohort revealed comparable virus-specific Ab responses in terms of binding levels, neutralization titers, and Fc-mediated activities of ADCP and complement deposition. However, the S2-specific Ig responses were distinguishing in that anti-S2 Ab levels tended to increase in hospitalized patients and decrease in ambulatory patients over time, and correlated with all other Ab parameters tested more strongly among hospitalized patients than ambulatory patients. Furthermore, the overall cytokine responses in sera, which were collected from four days to >7 months post-disease onset, were elevated in the hospitalized vs. ambulatory patients. The IL-4 and IL-27 levels were notably higher in the hospitalized group and showed no apparent decline over the >7-month period. The greater levels of IL-27, but not IL-4, were maintained when the hospitalized patients were matched to subsets of ambulatory patients by absence or presence of comorbidities, early or late sample collection time, and spike-specific total Ig levels. Nonetheless, the importance of S2-specific Abs and IL-27 in contributing to and/or resulting from severe COVID-19 remains unclear and warrants further investigation.

IL-27 is a heterodimeric IL-12 family cytokine, consisting of IL-12p35-related p28 and Epstein-Barr virus-induced 3 (EBI3) proteins (40). Together with IL-12, IL-27 is an initiator of Th1 polarization of CD4+ cells (41,42). IL-27 also promotes IFN-γ production by CD8+ T cells and NKT cells (43). Myeloid cell populations, including macrophages, inflammatory monocytes, microglia, and dendritic cells are the main sources of IL-27, which can be elicited by a range of microbial and immune stimuli (44). IL-27 signaling in turn can induce the release of pro-inflammatory cytokines such as IL-1, TNF-α, IL-18, and IL-12 (45,46). Indeed, our data showed that IL-27 levels correlated with those of IL-1α and TNF-α and that, with IL-27, IL-18 was elevated in hospitalized vs. ambulatory cases albeit without statistical significance. In a study looking at cytokine and leukocyte profile of 44 SARS-CoV-2-infected patients that were separated in two groups based on age (> or <60 years old), IL-27, together with CXCL8, IL-10, IL-15 and TNF-α, was associated to older age, longer hospitalization, and more severe COVID-19 (29). In contrast, in a Singapore cohort, no difference was observed in IL-4 and IL-27 levels between symptomatic and asymptomatic patients; rather, higher levels of MCP-1 and PDGF-BB were detected in patients with persistent symptoms vs. symptom-free patients (47). Beyond COVID-19, the levels of IL-27 were found to increase in patients with pulmonary inflammatory diseases including tuberculosis, asthma, influenza, acute lung injury, lung cancer, chronic obstructive pulmonary disease, acute lung injury, acute respiratory distress syndrome, and community-acquired pneumonia (48–53), indicating that an elevated IL-27 level is likely a common response to lung infections and injuries.

In contrast to Th1-promoting IL-27, IL-4 is a cytokine that induces Th2 differentiation. Upon activation by IL-4, Th2 cells subsequently produce additional IL-4 in a positive feedback loop. IL-4 is produced primarily by mast cells, Th2 cells, eosinophils and basophils (54). IL-4, along with other Th2 cytokines, is involved in the airway inflammation observed in the lungs of patients with allergic asthma (55). In our COVID-19 cohort, IL-4 levels were higher when the hospitalized cases were compared with all ambulatory patients but not with matched ambulatory patients. Conflicting results also have been published regarding IL-4, which was either associated with severe COVID-19 (19), or was thought to be beneficial for the recovery of COVID-19 patients (56).

A constellation of elevated cytokines has been associated with severe cases of COVID-19. Here we observed higher median values for several cytokines in hospitalized vs. ambulatory groups, although only IL-27 was significantly elevated in hospitalized patients as compared to all and matched ambulatory patients. Interestingly, in the hospitalized patients but not in the ambulatory patients, the levels of IL-27 correlated with those of IL-1α, IL-3, IL-6 and TNF-α, which were previously reported to be elevated and correlated with severe COVID-19 (28,29,57,58). A study looking at serum IL-6, IL-8 and TNF-α at the time of hospitalization also found that they were strong and independent predictors of patient survival in a cohort of hospitalized patients (57). Yet, another study measuring IL-6 and IL-18 serum levels found IL-18 to correlate with other inflammatory markers and reflect disease severity (59). And a different study looking at 48 cytokines, chemokines, and growth factors showed significantly higher IL-12 levels in the asymptomatic and mild disease groups than in the moderate and severe disease groups, while IL-4 levels were comparable and IL-27 was not examined (60). Nonetheless, these studies did not separate the individuals based on associated comorbidities, age, or racial/ethnic groups.

Limited information is available about cytokine responses among US minority groups, in particular African Americans and Hispanics who have higher rates of SARS-COV-2 infection, hospitalization, and death (30,61,62). African Americans are also more likely to have diabetes, hypertension, obesity, asthma, and heart disease, all of which are comorbidities associated with severe COVID-19. Moreover, little is known about cytokine levels in this population and in the context of associated comorbidities. The COVID-19-associated hospitalization rates are also higher among males than females (5.1 vs. 4.1 per 100,000) (61). Indeed, markers of brain and endothelial injury and inflammation were shown to be sex specifically regulated in SARS-CoV-2 infection (63). Nonetheless, scant information is available regarding the differences/similarities in cytokine levels between males and females, even though sex differences are likely to impact immune responses to SARS-CoV-2, as seen against other viruses, for examples, sex hormone-regulated pDC responses and sex differences in cytokine and chemokine production and neutrophil recruitment during influenza virus infection (64).

In terms of Ab responses, the levels of anti-SARS-CoV-2 Abs, in particular IgA, and neutralization activities have been positively associated with COVID-19 severity (8–10). A longitudinal study of Italian patients presenting a wide range of clinical manifestations identified anti-S1 IgA as an indicator of COVID-19 severity (65). We performed Ig isotyping with the entire spike protein, precluding assessment of Ig isotypes against S1 and S2 subunits. In another study, faster induction of S2-reactive IgG during the first week of infection, along with IgG cross-reactivity with the common human beta coronaviruses (β-hCoVs), correlated with COVID-19 severity, implicating a biased early response toward S2 epitopes cross-reactive with hCoVs (66). Comparison of spike proteins from SARS-CoV-2 (WA1) and hCoV strains shows varying levels of amino acid conservation across different spike regions. S1 and RBD exhibit only 25% to 30% identity, while S2 has higher percent identity: 41% to 42% with β-hCoVs (OC43 and HKU1), and 34% with α-hCoVs (229E and NL63). In our study, a distinguishable feature of Ab responses was the stronger concordance of Ig levels against S2 with anti-S1 Ig, anti-RBD Ig, and the other tested Ab parameters in the severe vs. ambulatory cases. These results suggest that the Ab responses to the S1 and S2 regions of spike were upregulated synchronously in the patients who went on to have severe COVID-19, but not in patients with mild cases. The S2-specific Ig responses were relatively low in both groups but tended to decline over time and disconnect from those of S1 in the ambulatory group. The S2 epitopes targeted by Abs from severe vs. mild cases in our study have not been determined, but Abs against certain sites in the more conserved S2 subunit may play a role in preventing or promoting progression to severe COVID-19. Neutralizing mAbs against S2 have been reported, such as 3A3 specific for a conserved epitope in the hinge region between the heptad repeat-1 helix and the central helix (67) and B1 recognizing the beta-hairpin region (68), although the prevalence of these Abs in COVID-19 patients is unknown. Garrido *et al*. demonstrated a correlation of IgG targeting different immunodominant conserved S2 regions with COVID-19 severity: Abs against heptad repeat-2 and S2’ fusion peptide correlated with milder and more severe disease, respectively (69); the functional aspects of these anti-S2 Abs are yet to be evaluated. Further studies also are warranted to focus on the activities of pre-existing and SARS-CoV-2-induced S2-specific Abs, including neutralization, Fab affinity, Fc glycosylation, and affinity for Fc receptors and complement, in patients with different disease severity and after COVID-19 vaccination.

Fc-mediated Ab functions also have been associated with COVID-19 severity. Abs-mediated FcγRIIa and FcγRIIIa activation positively correlated with symptom severity among ambulatory New York patients at 1-2 months post-disease onset (69). Additionally, IgG Fc glycome composition was shown to predict disease severity, with patients with a poor outcome having, at diagnosis, IgG deficient in galactosylation and sialylation and more bis-GlcNAc structures (13). Since the Fc glycan composition influences Fc-mediated functions, further investigation may reveal distinct Fc activities. We previously observed higher complement-binding potency of SARS-CoV-2 spike-specific Abs elicited by vaccination vs. infection (35). In this study, comparable complement binding potencies were seen in spike-specific Abs from hospitalized vs. ambulatory COVID-19 patients. ADCP capacity was also similar, while other Fc activities such as antibody-dependent cellular cytotoxicity and affinity for the different Fcγ receptors have not been studied.

The study was subject to additional limitations. The samples sizes were small, especially for the hospitalized patients, and this group included only survivors. Future study with larger sample sizes is needed to differentiate hospitalized patients requiring different levels of care and interventions and to match patient groups by race/ethic groups and specific comorbidities. In addition, all samples were collected from patients during the first wave of infection in New York City in March-December 2020, when the initial SARS-CoV-2 variant was predominant and no vaccine or antiviral therapeutic intervention were available. It is unknown if the data presented herein can be extrapolated directly to responses against Delta, Omicron, and other variants emerging in the future. Vaccines and therapeutics now widely available would also impact Ab and cytokine/chemokine responses to infection. The antiviral functional assays were also limited to neutralization, ADCP, and complement binding, and to the evaluation of the original SARS-CoV-2 variant using recombinant protein or pseudovirus. Lastly, our study is primarily cross-sectional with samples from different subjects over a wide range of time points post-infection. Although minimal changes were apparent in the levels of antibodies and cytokines during the observation period, the individual responses at the early and late time points may differ and were not studied herein. The longitudinal samples tested for the initial nucleoprotein-IgG study were unavailable for the subsequent experiments.

In summary, among the older and mostly male VA patients studied, SARS-CoV-2-specific Ab levels and functional activities did not distinguish hospitalized and ambulatory COVID-19 cases. However, a discordant S2-specific Ab response was noted among the ambulatory patients. Moreover, higher levels of cytokines, notably IL-4 and IL-27, were induced and maintained in hospitalized vs. ambulatory cases. These data offer a pathway to pursue for a better understanding of the immune mechanisms that play a role in protection against vs. progression to severe COVID-19.

## Data Availability

All data produced in the present study are available upon reasonable written request to the authors.

## Contributors

J.K., S.Z-P., C.U., and C.E.H. wrote and edited the manuscript. J.K., C.E.H., and S.Z-P. designed the experiments. J.K. and G.S.L. performed the experiments and collected the data. J.K., G.S.L, A.N., C.U. and C.E.H. analyzed the data. K.Y.O., F.A., and B.L. provided protocols, antigens, cells, and pseudovirus stocks. V.S. and the PARIS study group provided banked human samples, J.C.B., R.E-G., and J.J.I. obtained specimens. All authors read and approved the final manuscript.

PARIS Study Team: Angela A. Amoako, Dalles Andre, Mahmoud H. Awawda, Katherine F. Beach, Maria C. Bermúdez-González, Rachel L Chernet, Emily D. Ferreri, Daniel L. Floda, Joshua Hamburger, Giulio Kleiner, Neko Lyttle, Wanni A Mendez, Lubbertus CF Mulder, Ismail Nabeel, Kayla Russo, Ashley Beathrese T. Salimbangon, Miti Saksena, Levy A. Sominsky, Ania Wajnberg.

## Data sharing

Final de-identified data sets underlying this publication will be shared upon written request.

## Acknowledgments

We thank Dr. Florian Krammer for providing spike and RBD antigens, and all the study participants for their contribution to the research. We would like to thank the expertise and assistance of Dr. Christopher Bare and the Dean’s Flow Cytometry CORE at Mount Sinai.

## Figure legends

**Supplemental Fig 1. Total Ig antibody responses to SARS-CoV-2 among hospitalized and ambulatory VA patients**.

**a**. Total Ig levels against RBD, S1 and S2 in hospitalized (n = 10) vs. ambulatory (n = 42) patients.

AUC: area under the titration curve. The red line represents the median. The dotted line represents the cut-off, calculated as mean + 3SD of 5 COVID-19-negative specimens.

**b**. Total Ig levels against RBD, S1 and S2 between patients with (n = 24) vs. without (n = 28) comorbidities.

The red line represents the median. The dotted line represents the cut-off, calculated as mean + 3SD of 5 COVID-19-negative specimens.

**c**. Changes of RBD-, S1-, S2-, and nucleoprotein-specific total Ig levels over time post-symptom onset.

The dotted lines represent the linear regression 95% confidence bands.

**Supplemental Fig 2. RBD-specific antibody isotypes in sera of matched hospitalized and ambulatory VA patients**.

RBD-specific IgM, IgG1-4, IgA1 and IgA2 levels were measured in sera from matched hospitalized vs. ambulatory patients.

The data are presented as mean fluorescent intensity (MFI) at a 1:200 serum dilution. Each sample was tested in duplicate. The dotted line represents the cut-off.

Statistical significance was calculated by a Wilcoxon matched-pairs test; each hospitalized patient was compared to 2 or 3 matched ambulatory patients.

**Supplemental Fig 3. Fc-mediated activities of RBD-specific antibodies in sera from matched hospitalized and ambulatory VA patients**.

**a**. C1q binding to RBD-specific antibodies in matched hospitalized vs. ambulatory patients.

**b**. C3b binding to RBD-specific antibodies in matched hospitalized vs. ambulatory patients.

AUC: area under the titration curve. Statistical significance was calculated by a Wilcoxon matched-pairs test: each hospitalized patient was compared to 2 or 3 matched specimens from ambulatory patients.

**Supplemental Fig 4. Differential cytokine/chemokine responses in sera of COVID-19-positive vs. -negative VA patients**.

Forty-eight cytokines and chemokines were measured in serum specimens from COVID-19-positive (n = 52) vs. negative (n = 28).

Data are shown as concentration in pg/mL (log10). Red line represents median. Highlighted red boxes show cytokines or chemokines with significant differences (*: p<0.05, **: p<0.01, ***: p<0.001, ****: p<0.0001), calculated by unpaired non-parametric t-test (GraphPad Prism). Significance was similarly achieved with parametric t-test and non-parametric Mann Whitney Wilcoxon in R.

**Supplemental Fig 5. Levels of cytokines and chemokines in sera of COVID-19 VA patients over time**.

Changes of pg/mL concentrations of 11 cytokines (shown in Fig 5 to be statistically different between COVID-19-positive and –negative individuals) + IL-4 and IL-27 over time post-symptom onset.

The dotted lines represent the linear regression 95% confidence bands.

**Supplemental Fig 6. Differences in the levels of cytokines and chemokines among hospitalized and ambulatory VA patients**.

The levels of cytokines/chemokines were measured in sera from hospitalized (n = 10) vs. ambulatory (n =4 2) patients.

**a**. The data are shown as pg/mL concentration in log10. The gray line represents the median. The highlighted red boxes show the cytokines with significant differences between two patient groups (*: p<0.05, **: p<0.01), calculated by unpaired non-parametric t-test (GraphPad Prism). Significance was similarly achieved with parametric t-test and non-parametric Mann Whitney Wilcoxon in R.

**b**. The data are shown as median modified Z scores. p value was obtained from one-side t test comparing median values of all cytokines/chemokines in hospitalized vs. ambulatory groups.

**Supplemental Fig 7. Correlations of cytokines and chemokines in hospitalized, ambulatory, and control patients**.

Spearman correlation matrix of serum cytokines/chemokines levels were generated to compare hospitalized patients (n = 10), ambulatory patients (n = 42), and COVID-19-negative individuals (n = 28).

