## Supplemental Figures 1-7 and Tables 1-2 for "IMMUNE PROFILES TO DISTINGUISH HOSPITALIZED VERSUS AMBULATORY COVID-19 CASES IN OLDER PATIENTS"

### Suppl Figure 1

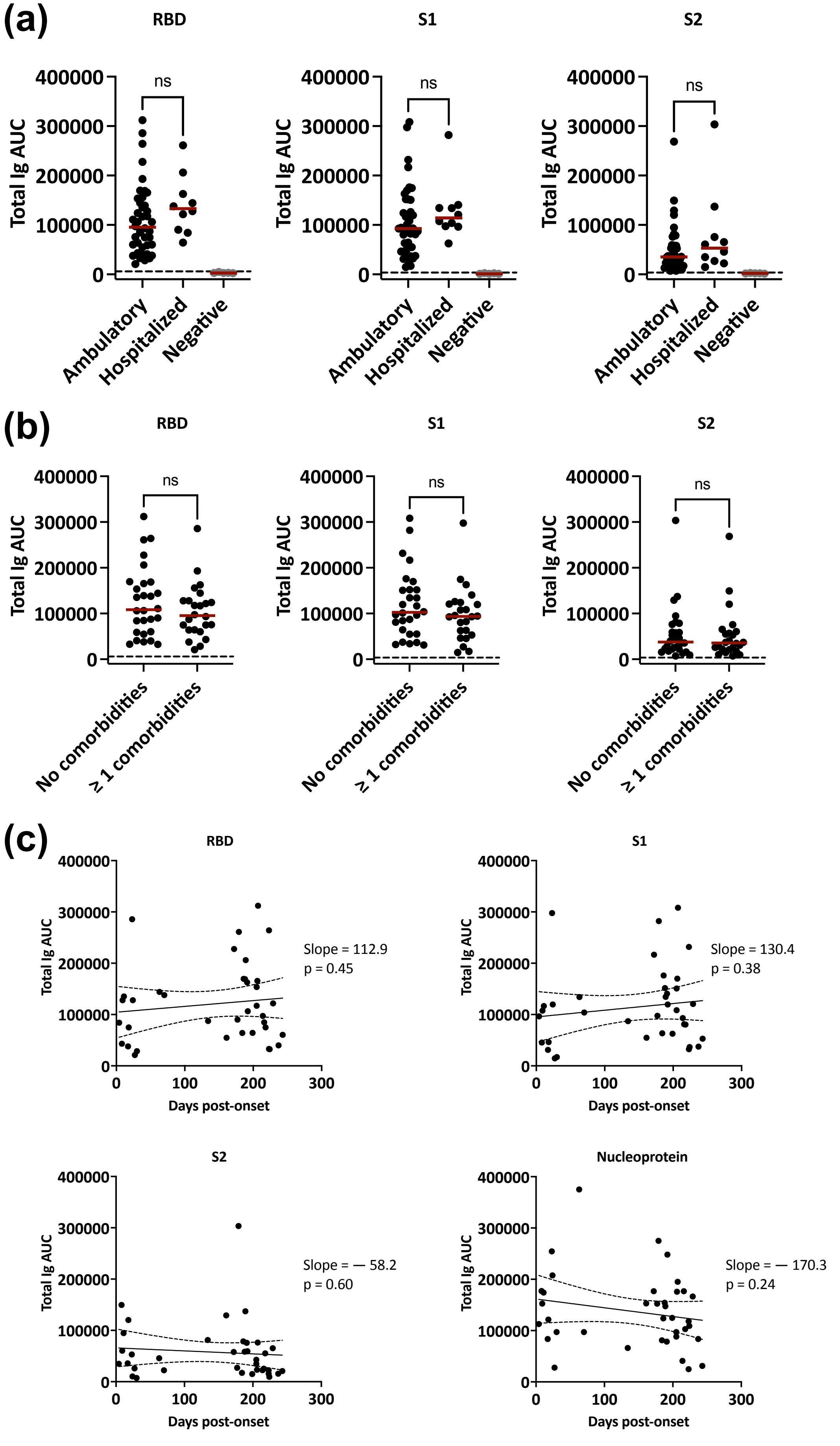

### Suppl Figure 2

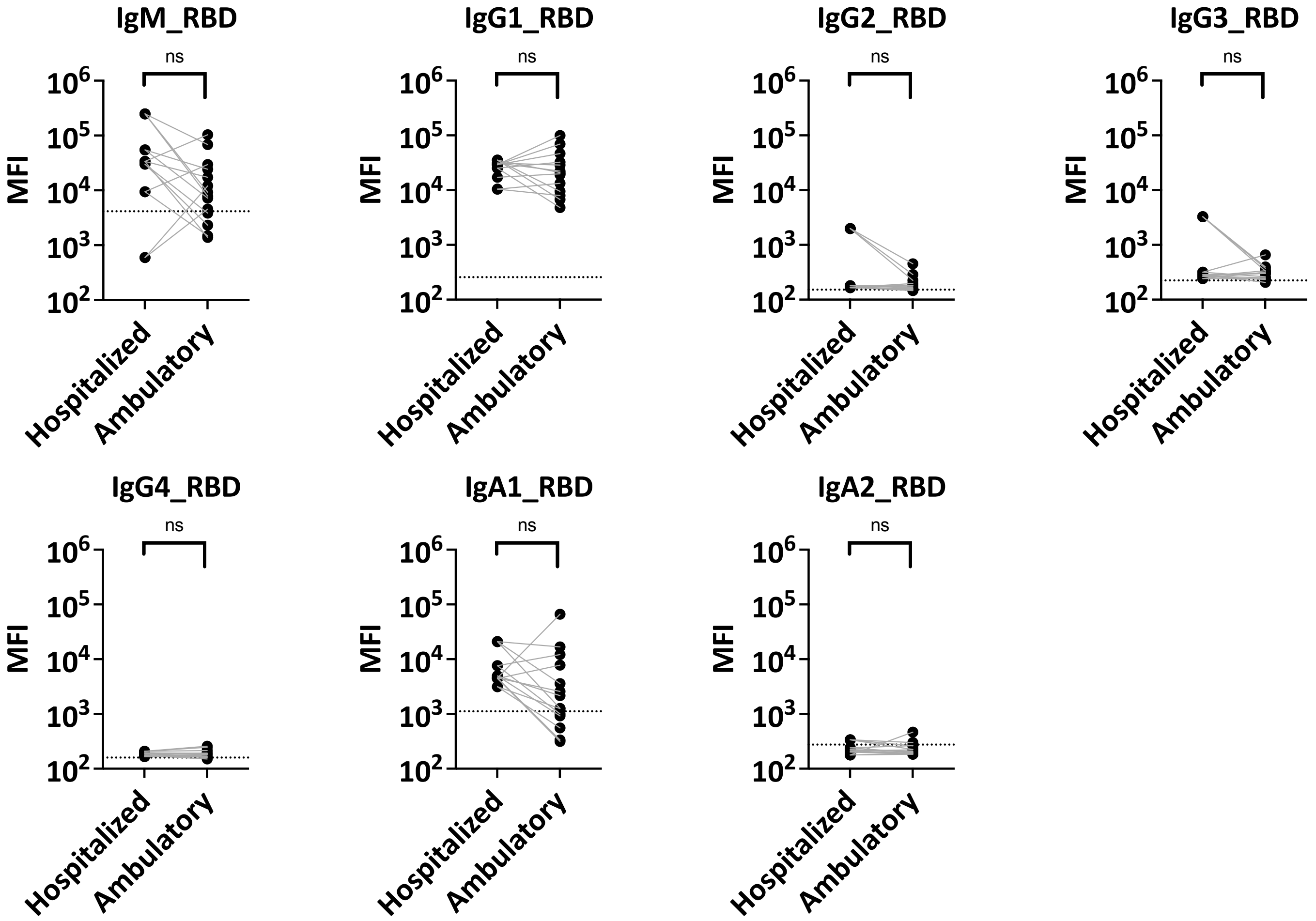

### Suppl Figure 3

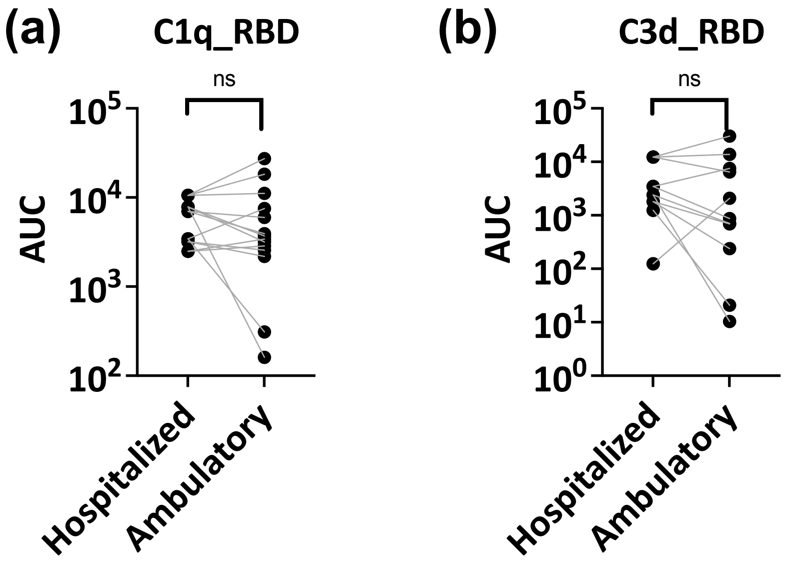

### Suppl Figure 4

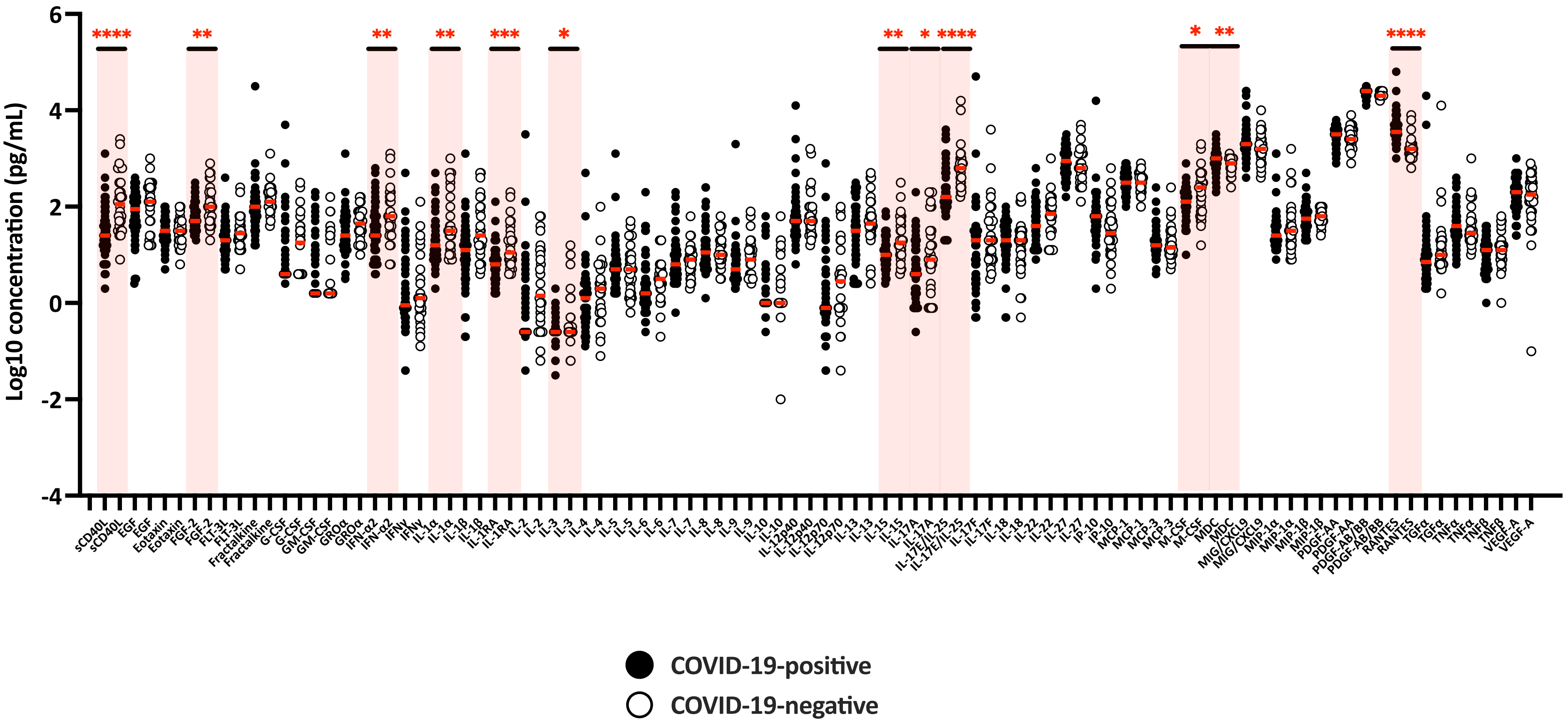

### Suppl Figure 5

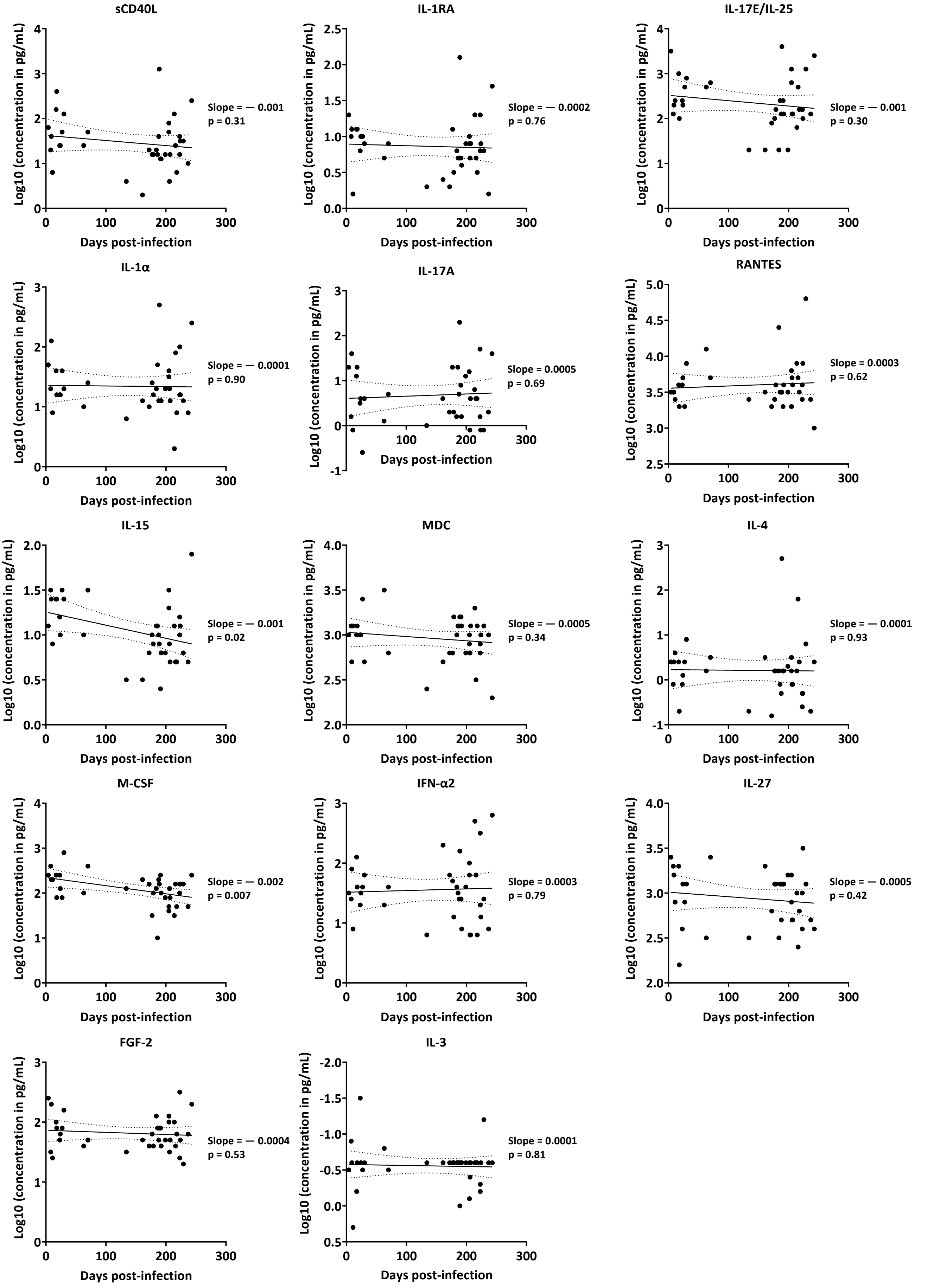

### Suppl Figure 6

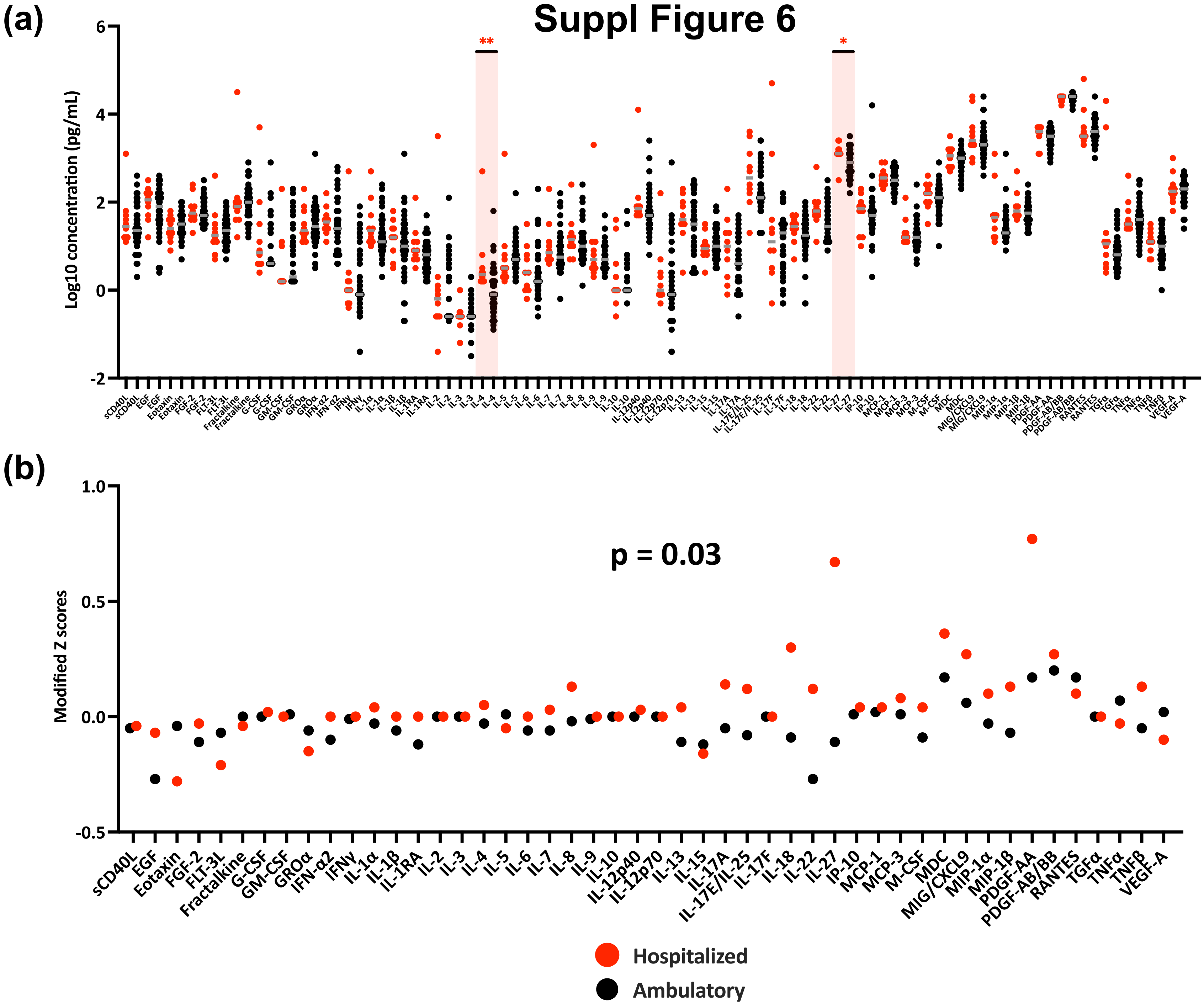

### Suppl Figure 7

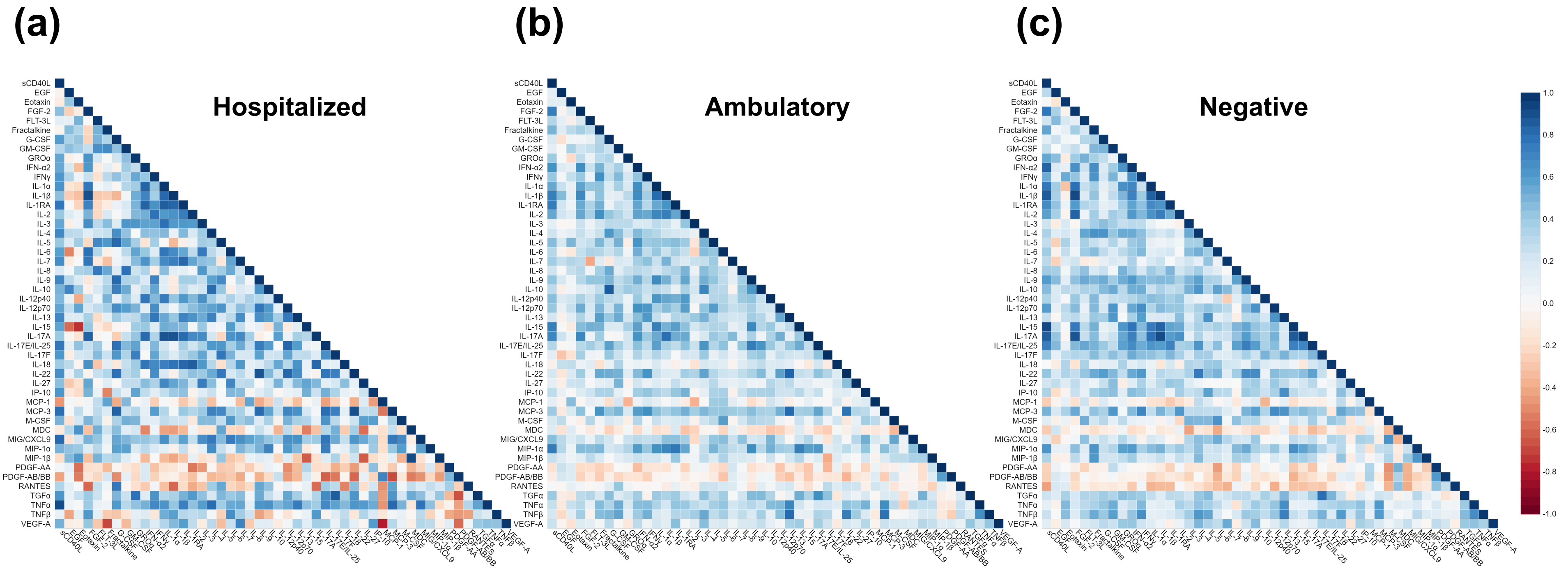

**Supplemental Table 1: Clinical data of all COVID-19-infected patients in the VA cohort (n = 52).**

| Code | Age | Sex | Clinical symptoms | Severity | Comorbidities | Race | Ethnicity | Serum collection (Days post-infection) |
|---|---|---|---|---|---|---|---|---|
| **CVAP1** | 40-44 | M | Bilateral pneumonia | Hospitalized | | White | Hispanic | 177 |
| **CVAP2** | 75-79 | M | | Ambulatory | | Black | | 11 |
| **CVAP3** | 70-74 | M | Pneumonia | Hospitalized | | Black | | 189 |
| **CVAP4** | 60-64 | M | | Ambulatory | | White | Hispanic | 172 |
| **CVAP6** | 70-74 | M | | Ambulatory | | Black | | 192 |
| **CVAP7** | 80-84 | M | | Ambulatory | | White | Hispanic | 17 |
| **CVAP8** | 30-34 | M | | Ambulatory | | White | Hispanic | 224 |
| **CVAP9** | 90-94 | M | Pneumonia | Hospitalized | | Black | Hispanic | 179 |
| **CVAP10** | 50-54 | F | | Ambulatory | | Black | Hispanic | 186 |
| **CVAP12** | 40-44 | F | | Ambulatory | | Black | | 188 |
| **CVAP13** | 50-54 | M | | Ambulatory | | White | Hispanic | 206 |
| **CVAP14** | 55-59 | M | | Ambulatory | | White | Hispanic | 207 |
| **CVAP15** | 45-49 | M | | Hospitalized | | Black | | 63 |
| **CVAP16** | 55-59 | M | Bilateral pneumonia | Hospitalized | | White | Hispanic | 70 |
| **CVAP17** | 80-84 | M | Gastrointestin al bleeding | Hospitalized | | Black | | 4 |
| **CVAP18** | 75-79 | M | | Ambulatory | | White | | / |
| **CVAP19** | 55-59 | M | | Ambulatory | | White | Hispanic | 161 |
| **CVAP20** | 65-69 | M | | Ambulatory | Obesity, asthma | Black | | 218 |
| **CVAP21** | 45-49 | F | | Ambulatory | | Black | | / |
| **CVAP22** | 35-39 | M | | Ambulatory | Obesity | White | Hispanic | / |
| **CVAP23** | 70-74 | M | Bilateral pneumonia | Hospitalized | Chronic obstructive pulmonary disease (COPD) | Black | | 229 |
| **CVAP24** | 75-79 | M | | Ambulatory | | White | | 216 |
| **CVAP25** | 65-69 | M | | Ambulatory | Asthma, hypertension | White | Hispanic | / |
| **CVAP26** | 70-74 | M | Mild infiltration | Hospitalized | Coronary artery disease (CAD) | White | | 191 |
| **CVAP27** | 60-64 | M | | Ambulatory | Prostate cancer | White | | / |
| **CVAP28** | 60-64 | M | | Ambulatory | Chronic obstructive pulmonary disease (COPD), emphysema | Black | Hispanic | / |
| **CVAP29** | 70-74 | M | | Ambulatory | Hypertension | White | Hispanic | / |
| **CVAP30** | 55-59 | M | | Ambulatory | Obesity, hypertension | Black | | / |
| **CVAP31** | 65-69 | M | | Ambulatory | Asthma, hypertension | Black | | 214 |
| **CVAP32** | 55-59 | M | | Ambulatory | Obesity | Black | | 184 |

| | | | | | | | | |
|---|---|---|---|---|---|---|---|---|
| **CVAP33** | 75-79 | M | | Ambulatory | | White | Hispanic | / |
| **CVAP34** | 70-74 | M | | Ambulatory | Hypertension | White | Hispanic | / |
| **CVAP35** | 35-39 | F | | Ambulatory | | White | | / |
| **CVAP36** | 75-79 | M | | Ambulatory | Diabetes mellitus (DM) | Black | Hispanic | / |
| **CVAP37** | 65-69 | M | | Ambulatory | HIV | Black | | / |
| **CVAP38** | 75-79 | M | | Ambulatory | Diabetes mellitus (DM), coronary artery disease (CAD) | White | | 205 |
| **CVAP39** | 50-54 | F | | Ambulatory | | Black | | 205 |
| **CVAP40** | 40-44 | M | Bilateral pneumonia | Hospitalized | Diabetes mellitus, obesity | White | Hispanic | 199 |
| **CVAP41** | 55-59 | M | | Ambulatory | | Black | | 134 |
| **CVAP42** | 75-79 | M | | Ambulatory | Prostate cancer, hypertension | White | Hispanic | 24 |
| **CVAP43** | 55-59 | F | | Ambulatory | | Black | | 237 |
| **CVAP44** | 55-59 | M | | Ambulatory | | Black | | / |
| **CVAP45** | 40-44 | F | | Ambulatory | | Black | | 223 |
| **CVAP46** | 55-59 | F | | Ambulatory | | Black | | 223 |
| **CVAP47** | 80-84 | M | | Ambulatory | Prostate cancer, hypertension | Black | Hispanic | 243 |
| **CVAP49** | 60-64 | M | Bilateral pneumonia | Hospitalized | Pulmonary sarcoidosis, asthma | | | 9 |
| **CVAP50** | 35-39 | F | | Ambulatory | | White | Hispanic | / |
| **CVAP51** | 65-69 | M | | Ambulatory | End stage renal disease (ESRD) | White | | 30 |
| **CVAP52** | 45-49 | F | | Ambulatory | Ulcerative colitis | Black | | 27 |
| **CVAP53** | 60-64 | M | | Ambulatory | HIV, coronary artery disease (CAD), diabetes mellitus (DM) | White | | 23 |
| **CVAP54** | 70-74 | M | | Ambulatory | End stage renal disease (ESRD) | Black | | 8 |
| **CVAP55** | 80-84 | M | | Ambulatory | Chronic obstructive pulmonary disease (COPD), pulmonary embolism (PE) | Black | Hispanic | 18 |

/: no data available and excluded in Figure 1E.

**Supplemental Table 2. Clinical data of hospitalized and ambulatory COVID-19 patients matched for sex, days post-infection, total Ig and comorbidities (six sets, n = 20 total).**

| | Code | Sex | Days post-infection | Comorbidities | Spike-specific total Ig AUC | Severity | Clinical symptoms |
|---|---|---|---|---|---|---|---|
| Set 1 | CVAP1 | M | 177 | No | 80083 | Hospitalized | Bilateral Interstitial Pneumonia |
| | CVAP24 | M | 216 | No | 71073 | Ambulatory | |
| | CVAP41 | M | 134 | No | 101739 | Ambulatory | |
| Set 2 | CVAP3 | M | 189 | No | 257621 | Hospitalized | Pneumonia |
| | CVAP4 | M | 172 | No | 212313 | Ambulatory | |
| | CVAP13 | M | 206 | No | 184248 | Ambulatory | |
| | CVAP14 | M | 207 | No | 284795 | Ambulatory | |
| Set 3 | CVAP17 | M | 4 | No | 79206 | Hospitalized | Gastrointestinal bleeding |
| | CVAP2 | M | 11 | No | 132416 | Ambulatory | |
| | CVAP7 | M | 17 | No | 48953 | Ambulatory | |
| Set 4 | CVAP23 | M | 229 | Yes | 157024 | Hospitalized | Bilateral Interstitial Pneumonia |
| | CVAP31 | M | 214 | Yes | 82607 | Ambulatory | |
| | CVAP38 | M | 205 | Yes | 120391 | Ambulatory | |
| Set 5 | CVAP40 | M | 199 | Yes | 56680 | Hospitalized | Bilateral Interstitial Pneumonia |
| | CVAP20 | M | 218 | Yes | 87902 | Ambulatory | |
| | CVAP32 | M | 184 | Yes | 83676 | Ambulatory | |
| Set 6 | CVAP49 | M | 9 | Yes | 127891 | Hospitalized | Bilateral Interstitial Pneumonia |
| | CVAP42 | M | 24 | Yes | 172417 | Ambulatory | |
| | CVAP54 | M | 8 | Yes | 88384 | Ambulatory | |
| | CVAP55 | M | 18 | Yes | 71418 | Ambulatory | |
